# Randomized Phase 2 Trial of Lirilumab as maintenance Treatment in Acute Myeloid Leukemia: Results of the EFFIKIR Trial

**DOI:** 10.1101/2024.08.23.24312477

**Authors:** N. Vey, AS. Chretien, PY. Dumas, C. Recher, L. Gastaud, B. Lioure, CE. Bulabois, C. Pautas, JP. Marolleau, S. Lepretre, E. Raffoux, X. Thomas, Y. Hichri, C. Bonmati, B. Quesnel, P. Rousselot, E. Jourdan, JV. Malfuson, G. Guillerm, JH. Bourhis, M. Ojeda-Uribe, M. Hunault, A. Ben Amara, MS. Rouvière, N. Boucherit, P. André, C. Preudhomme, N. Dulphy, A. Toubert, N. Ifrah, D. Olive, H. Dombret

## Abstract

Lirilumab is a fully human monoclonal antibody designed to block killer inhibitory receptors (KIR), which are major immune checkpoints involved in the regulation of NK cell-mediated killing of HLA-I-expressing tumors. EFFIKIR is a multicenter randomized double-blind 3-arm placebo-controlled phase II trial with lirilumab as single-agent as maintenance therapy of elderly patients with AML in first complete remission (NCT01687387). Two dose schedules led to either continuous or intermittent KIR occupancy. 153 patients were randomized and 152 patients were treated after 3+7 induction therapy. The median follow-up was 36.6 months. Lirilumab was well tolerated, with no significant hematological toxicity. The median LFS were 17.6, 6.7 and 13.9 months in the 0.1mg/kg arm, 1mg/kg arm and placebo arm, respectively. An excess in early relapse led to early termination of treatment in the 1mg/kg arm. Extensive analysis of immune cell fate following KIR blockade evidenced a decrease of KIR^+^ NK cell absolute counts following KIR blockade, associated with a decrease of Bcl-2. Lirilumab also bound antigen-experienced CD8^+^ T cells, and induced a transient decrease of CD69 expression. Besides, lirilumab bound vδ2^+^ γδT cells with a high cytotoxic potential, and induced a decrease of DNAM-1 and Bcl-2, the latter being associated with a decrease of KIR^+^ γδT cell, and with a drastic reduction of time to relapse. Overall, the potentially deleterious effects on immune effectors may have resulted in the impairment of immune surveillance associated with an unexpected high rate of early relapse in the group of patients exposed to prolonged full KIR blockade.

**KEY POINTS:**

- Prolonged full KIR blockade leads to potentially deleterious effects on NK cells, CD8^+^ T cells and vδ2^+^ γδT cells
- Combined inhibitory effects of KIR blockade may have resulted in the impairment of immunosurveillance associated with high rate of relapse

## INTRODUCTION

Killer cell Immunoglobulin-like Receptors (KIRs) are a class of major immune checkpoints that negatively regulate Natural Killer (NK) cell anti-tumor activity.^1,2^ Inhibitory KIR, KIR2DL and KIR3DL, display extracellular immunoglobulin-like domains conferring specificity for HLA-C or HLA-A/B allotypes, respectively. Blockade of inhibitory NK receptors has therefore the unique potential to enhance tumor cell killing without affecting healthy tissues. NK cells are cytotoxic against malignant cells, including Acute Myeloid Leukemia (AML)^1,3^, and AML-induced NK cell alterations are associated with a poor prognosis.^4,5^ KIR blockade by IPH2101, an anti-KIR monoclonal antibody, was found to be generally safe and displayed signals of activity in hematologic malignancies.^6^ Lirilumab (IPH2102/BMS-986015/BMS-986015-01) is a fully human IgG4 monoclonal antibody that binds specifically and with high affinity to the main human inhibitory KIR, KIR2DL1 and KIR2DL2/3. Lirilumab binding blocks the interaction of KIR2DL with HLA-C allotypes, prevents inhibitory signals, and fosters activation NK cell activation. In a phase I study of lirilumab in patients with hematological malignancies and solid tumors, we have shown that as for IPH2101, lirilumab was safe and well tolerated.^7^

AML in older patients is associated with a dismal prognosis. With conventional intensive chemotherapy, complete remission (CR) rates are comprised between 50% and 70% and the relapse rate is high after remission, which results in median leukemia-free survival (LFS) of about 12 months, median overall survival (OS) of 9 months and only 5-10% of patients are long-term survivors.^8–12^ There is thus a need to develop effective maintenance strategies, and immunotherapies targeting NK cell represent attractive approaches.

In our previous phase I study of lirilumab^7^, we observed that different pharmacodynamic profiles where associated with lirilumab dose: a transient saturation with the lowest dose versus a sustained full saturation with higher doses (0.015mg/kg versus 0.3mg/kg and above in that study). Evidence has highlighted the importance of KIR receptors in NK cell maturation and education^13,14^ and suggests that, in the clinic, transient KIR blockade might allow both the optimal activation of NK cells and the education of new NK cells towards the end of each treatment cycle when occupancy is reduced.

In order to evaluate the clinical efficacy of lirilumab and to determine the optimal dose-schedule, we designed a 3-arm phase II placebo-controlled study which compared the LFS of elderly patients in first CR of AML treated with two dose-schedules of lirilumab or placebo. A subset participated to a correlative study allowing for the double-blinded evaluation of the functional and phenotypic changes induced by the 2 treatment arms compared to the control group. The endpoints of the study were not met, and excess in early relapse led to early termination of the study. Extensive analysis of the immune cell fate following KIR blockade provides key elements to elucidate the potential mechanisms that led to this unexpected excess of relapse despite inhibition of a major immune checkpoint.

## PATIENTS AND METHODS

### Study design and treatment

This is a prospective multicenter double-blinded phase II trial comparing two dose-schedules of Lirilumab to a placebo control administered in the maintenance therapy of elderly patients with AML in first CR following intensive chemotherapy. The study was conducted in 41 centers of the French ALFA and FILO cooperative groups.

Patients were randomized to one of the two treatment arms or placebo in a 1:1:1 ratio, with minimization performed according to center, type of AML (primary versus secondary), number of consolidation cycles (one versus two), and cytogenetics (high versus intermediate risk). Lirilumab was administered at 0.1mg/kg every 12 weeks (arm A) or at 1mg/kg every 4 weeks (arm B) as a 1-hour IV infusion. Placebo (normal saline) was to be administered every 4 weeks as a 1-hour IV infusion in the placebo arm (arm C) and on weeks with no active treatment in arm C, to ensure blinding. Patients were scheduled to receive 28-day cycles for up to 24 months in the absence of intolerance or disease relapse. The study scheme is depicted in Fig. 1. Lirilumab was provided by Innate Pharma (Marseille, France) as 5 mL vials containing 10 mg/mL of lirilumab containing solution.

**Figure 1:**
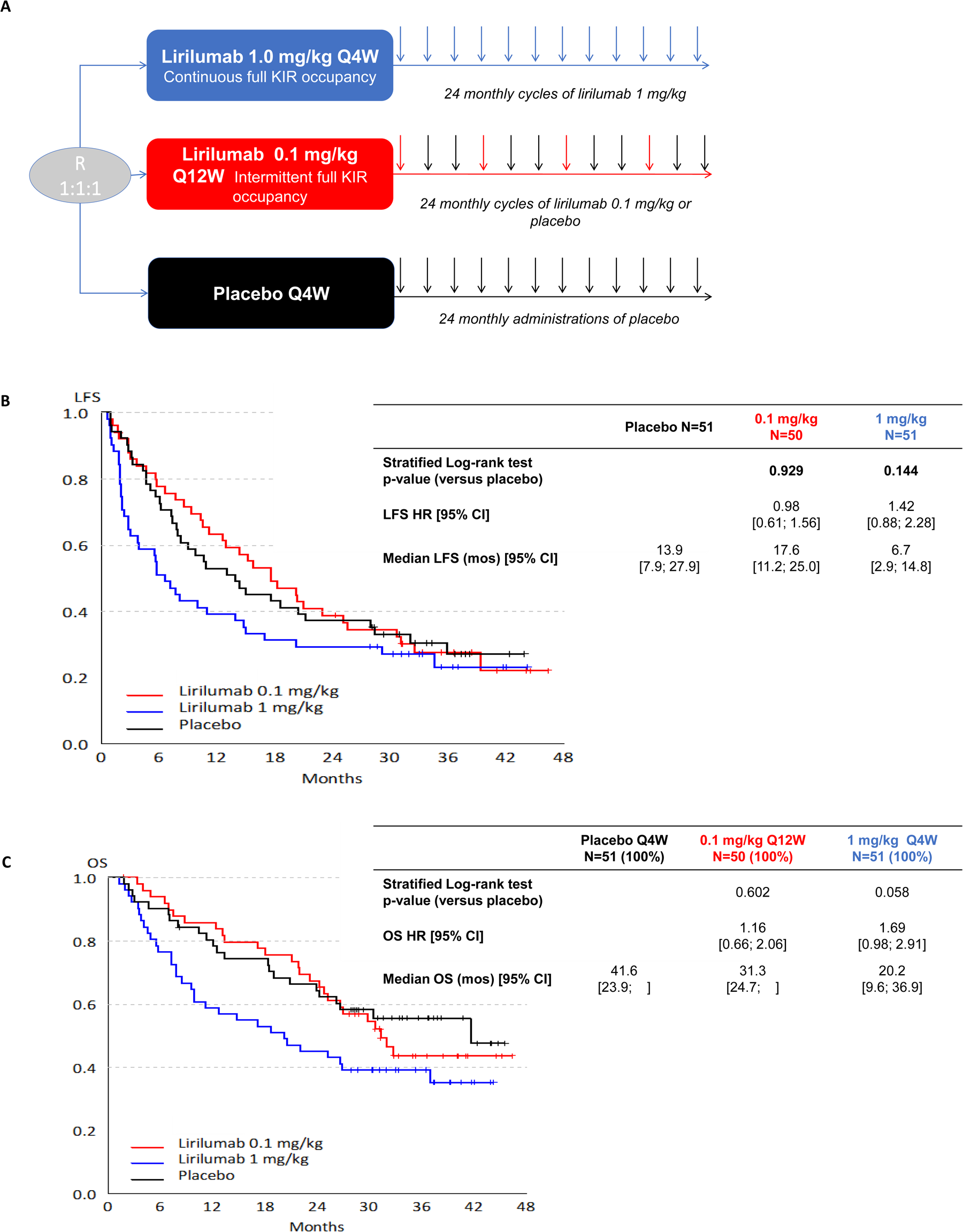
Clinical outcome in patients treated with lirilumab 1.0 mg/kg, lirilumab 0.1 mg/kg or placebo. A: Study design. B: Primary endpoint: Leukemia-Free Survival. C: Secondary endpoint : Overall Survival.

Study was approved by a national ethics committee (CPPRB) and registered at clinical trial.gov (NCT01687387).

### Patients

Main study eligibility criteria were: patients with de novo or secondary AML defined according to World Health Organization 2008 criteria^8^, in first CR or CRi (according to the revised recommendations of the International Working Group^15^) achieved after one or two conventional induction chemotherapy and who had received one or two consolidation cycles. Two different anthracycline-cytarabine containing induction and consolidation regimens described in the Supplementary data section had been used as recommended by each cooperative group. Induction chemotherapy had to have been administered within 6 months before randomization and consolidation within 3 months following CR. Patient had to be non-eligible for an allogeneic hematopoietic stem cell transplantation (allo-HSCT), to be aged of 60 to 80 years, to have an Eastern Cooperative Oncology Group (ECOG) performance status of 0 or 1; to have adequate renal and hepatic function. Patients with a history of auto-immune disease, patients on steroids were not eligible. All patients gave written informed consent prior to inclusion in the study.

### Efficacy and safety assessments

Safety was to be assessed, using Common Terminology Criteria for Adverse Events (CTCAE) version 4.03. A safety analysis was performed every 6 months for safety assessment by the DSMB. The primary efficacy variable for this trial was LFS, defined as the time elapsed between randomization and the occurrence of disease relapse or death from any cause. Secondary efficacy variables were the time to relapse (TTR), OS. TTR was defined as the time elapsed between randomization and the occurrence of disease relapse; OS was the time between randomization and death from any cause. Patients who had not progressed or died by the time of the final analyses, were censored as of the date of last contact for calculation of LFS, OS and TTR. Assessment of disease relapse was performed every 4 weeks during the treatment and during follow-up periods. Relapse was defined according to IWG criteria.^15^

### Statistical analysis

The planned sample size for the study comprised 150 patients (i.e., 50 patients were to be randomized to each treatment arm). This sample size was estimated in order to demonstrate a hazard ratio (HR) for LFS of 0.6 in favor of maintenance therapy with an 80% power for a one-sided log-rank test at an overall alpha level of 5%. The final sample size consisted of 169 screened patients, 153 randomized patients, and 152 treated patients. All efficacy analyses were performed using data from the Intention-to-treat (ITT) Population, which consisted of all subjects who received at least one dose of study medication. The Safety Population used for the safety analyses consisted of all patients who received at least one dose of lirilumab.

Following a DSMB meeting held on 10 March 2015, a unanimous recommendation was made to stop further treatment for ongoing patients treated with lirilumab 1mg/kg due to an apparent excess of early relapses compared with the two other groups. It was decided to keep the same assumptions regarding the number of LFS events to be observed in these two groups, i.e. 69 LFS events. Consequently, the primary efficacy analysis was performed on these two groups, with the alpha level decreased to 2.5% and the power to 55%.

LFS was analyzed using the log-rank test stratified by the following stratification factors, as recorded in the randomization file: primary versus secondary AML, and cytogenetics (high versus intermediate risk). The stratification factor “number of consolidation cycles (1 versus 2)” was not used in the log-rank test because there were too few patients in some combinations of strata and the log-rank test failed in this situation.

### Mass cytometry

PBMC were collected at C1D1H0, C1D8, C4D1, C7D1 and end of study (EOS), and cryopreserved in 10% DMSO. Forty-three patients participated to this correlative study (12 in arm 0.1mg/kg, 16 in arm 1.0mg/kg and 15 in the control arm. A total of 141 samples were analyzed among which 39 were collected at C1D1H0, 35 at C1D8, 26 at C4D1, 18 at C7D1, and 23 at EOS. PBMCs were processed as previously described.^5,16^ The detailed protocol is provided in appendix. Lymphocytes were exported for further analysis using opt-SNE^17^. Lirilumab-bound cells were identified using a metal-conjugated anti-IgG4 antibody.

## RESULTS

### Patient characteristics

Between November 2012 and July 2014, 169 patients were screened, 153 were randomized, and 152 were treated (Table 1). One patient was not treated in the lirilumab 0.1mg/kg arm, due to consent withdrawal before treatment initiation. The final numbers of patients were 50 in the lirilumab 0.1mg/kg arm, 51 in the lirilumab 1mg/kg arm, and 51 in the placebo arm (Fig. 1A). No significant imbalances were seen across treatment arms. Median age was 70 years. One fourth of patients had a favorable European LeukemiaNet (ELN) 2010 genetic subtype of AML. The median time since AML diagnosis was 4.8 month and 81% of patients had received two consolidation cycles prior to inclusion.

**Table 1.** Patient characteristics.

|  | **1 mg/kg Q4W** | **0.1 mg/kg Q12W** | **Placebo Q4W** |
| --- | --- | --- | --- |
|  | **N=51 (100%)** | **N=50 (100%)** | **N=51 (100%)** |
| Median age years (range) | 70 (60-80) | 70 (60-80) | 69 (60-77) |
| Sex (Female) | 28 (55%) | 16 (32%) | 17 (33%) |
| ECOG |  |  |  |
| 0 | 28 (55%) | 27 (54%) | 35 (69%) |
| 1 | 23 (45%) | 23 (46%) | 16 (31%) |
| Secondary AML | 6 (12%) | 9 (18%) | 7 (14%) |
| ELN 2010 risk category |  |  |  |
| - Favorable | 15 (29%) | 12 (24%) | 12 (24%) |
| - Intermediate-1 / 2 | 25 (49%) | 27 (54%) | 26 (51%) |
| - Adverse | 5 (10%) | 6 (12%) | 5 (10%) |
| Prior therapy |  |  |  |
| - Induction (1 cycle / 2 cycles) | 50 (98%) / 1 (2%) | 50 (100%) / 0 | 48 (94%) / 3 (6%) |
| - Consolidation (1 cycle / 2 cycles) | 10 (20%) / 41 (80%) | 9 (18%) / 41 (82%) | 10 (20%) / 41 (80%) |
| - Consolidation (1+5 / IDAC) | 28 (55%) / 23 (45%) | 20 (40%) / 29 (58%) | 32 (63%) / 19 (37%) |
| Median months since CR (range) | 3.0 (1.1-5.0) | 3.3 (1.3-5.5) | 3.3 (2.0-5.9) |

### Treatment

Following a Data and Safety Monitoring Board (DSMB) recommendation on 10 March 2015, the decision was made to stop further treatment for ongoing patients treated with lirilumab 1mg/kg due to an apparent excess of early relapses compared with the two other groups. At that time, recruitment had already been completed. As a result, exposure was greater in the lirilumab 0.1mg/kg arm (median number of cycles of 16) and in the placebo arm (median number of cycles of 11) as compared to the lirilumab 1mg/kg arm (median number of cycles of 8).

Relapse (62.7%) was the main reason for discontinuation (52.9% from the 0.1mg/kg arm, 72.5% from the 1mg/kg arm, and 62.7% from the placebo arm), followed by adverse events (AEs) (10.5%), withdrawal of consent, investigator’s decision, and sponsor’s decision (1.3% each) (Fig. S1).

### Efficacy results

Efficacy analysis concerned only the comparison between lirilumab 0.1mg/kg versus placebo (Fig. 1). The study did not meet its primary endpoint, as no significant difference between lirilumab 0.1mg/kg and placebo was seen (stratified LFS Hazard ratio (HR): 0.98 (95% confidence interval [CI], 0.61 to 1.56, p=0.936). The median LFS were 17.6, 6.7 and 13.9 months in the 0.1mg/kg arm, 1mg/kg arm and placebo arm, respectively. Regarding the comparison between lirilumab 1mg/kg and placebo, no statistically significant difference was observed, but LFS in the lirilumab 1mg/kg arm was inferior to that in the placebo arm (median LFS of 6.7 months, HR 0.95; 95% CI, 0.59 to 1.53; p=0.177) (Fig. 1B).

With regard to secondary efficacy endpoints, no significant differences were found in any of the comparisons which TTR times, median OS (31.3, 20.2 and 41.6 months in the 0.1mg/kg arm, 1mg/kg arm and placebo arm, respectively, p=0.602 and 0.058) (Fig. 1C).

No prognostic factor for LFS for the comparison between lirilumab 0.1mg/kg and placebo was seen. For the comparison of lirilumab 1mg/kg with placebo, the type of induction therapy (idarubicin vs daunorubicin) and the number of platelets at the time of inclusion (<100 vs ≥100 x10^9^/L) were identified in the Cox model as independent prognostic factors for LFS (HR=1.35 and 4.14, respectively). Results are summarized as Forest Plots displayed in Fig. S2A and S2B.

As previously mentioned, an excess in early relapse was observed and justified early termination of treatment in the 1mg/kg arm. Although LFS was not statistically different between arms, curves clearly show an early drop in LFS in this arm as compared to the 2 others.

### Safety

Overall, 91.4% of patients had at least one treatment-emergent AE. The event rates (i.e. the number of patients with AEs divided by the total drug exposure in months), was lower in the lirilumab 0.1mg/kg arm than in the lirilumab 1mg/kg and the placebo arms. The most frequent AEs are presented in Table 2. Infusion-related reactions were the most frequent, did not reach a grade 4, while grade 3 reactions frequency was nearly identical in the three arms.

**Table 1.**
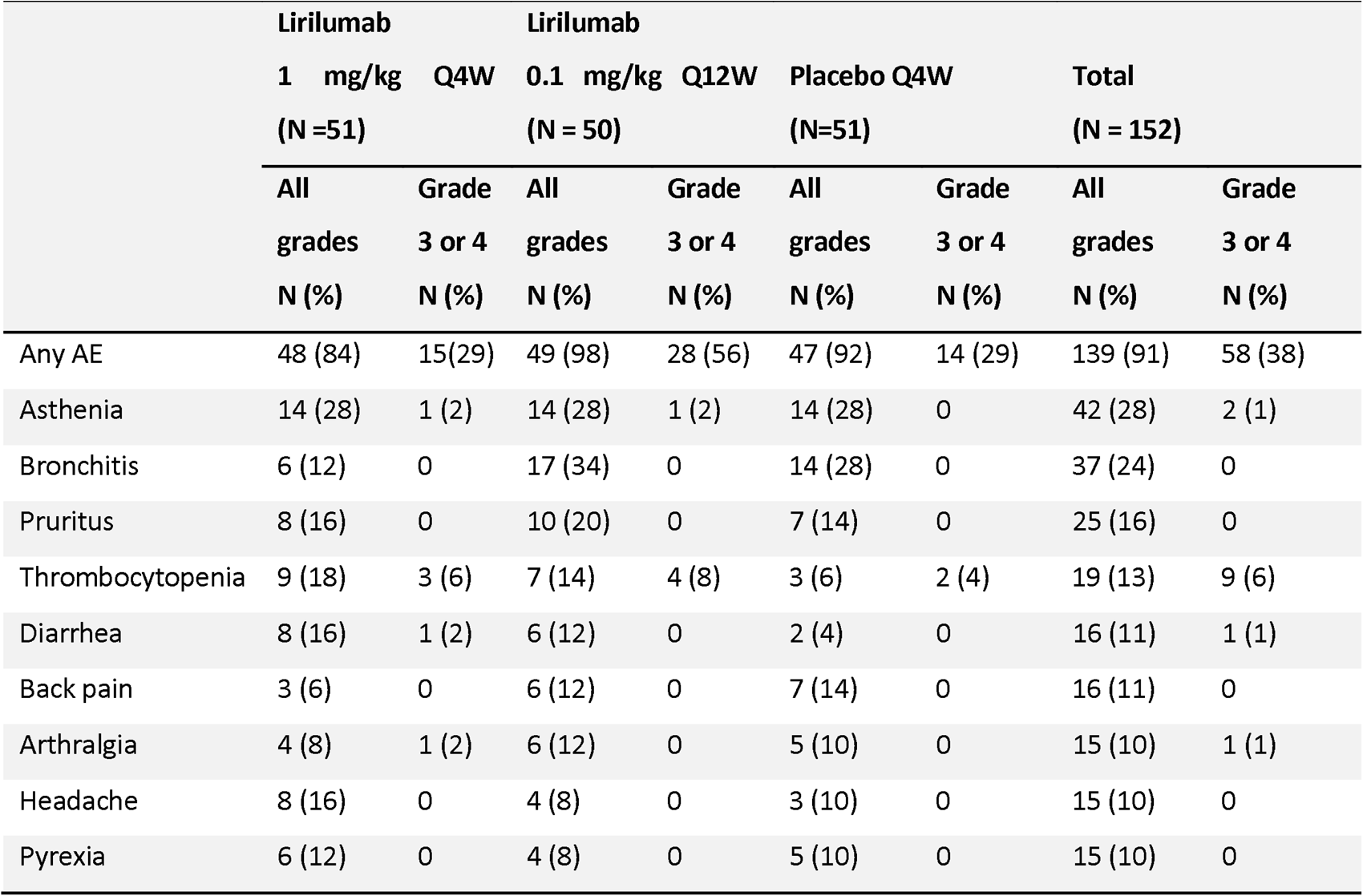
Adverse events occurring in <10% of patients.

No significant hematological toxicity was seen in the 3 arms. Eighteen malignant tumors were reported in 14 patients. 75% of patients were in the lirilumab 0.1mg/kg arm. There were 3 basal-cell carcinoma, and the other six neoplasms had each a maximum of one case. In the other two arms combined, there were 3 myeloid malignancies, 6 solid tumors (including 2 basal cell carcinomas).

A total of 81 deaths were reported during the study: 26 in the lirilumab 0.1mg/kg arm, 32 in the lirilumab 1mg/kg arm, and 23 in the placebo arm. Nearly 85% of such deaths were attributed to disease progression across the three arms.

### Pharmacokinetics and pharmacodynamics

PK-PD sampling was performed for the first 60 randomized patients. Intermittent full KIR occupancy was observed with the dose of 0.1mg/kg every 3 months, with full KIR occupancy lasting 7 days to 1 month for the majority of the patients. Long-term full KIR occupancy was observed at the dose of 1mg/kg every 4 weeks (Fig. S3).

### Immune modifications during treatment

Lirilumab was not immunogenic with 15 patients presenting Human Anti-Human Antibodies, all with low titers. Of note, 9 out of the 15 positive patients tested positive at baseline and 5/15 patients were in the placebo arm (data not shown). Lirilumab-induced immune modifications were then extensively analyzed by mass cytometry. For these experiments, we used a metal-conjugated anti-IgG4 antibody in order to specifically identify lirilumab-bound immune cells (Fig. 2A). As expected, based on prior *in vitro* studies^18^, lirilumab bound KIR-expressing immune populations, including NK cells, Vγ9Vδ2^+^ γδT cells, CD8^+^ T cells, and to a lesser extent CD4^+^ T cells (Fig. 2B-C). Eight days after lirilumab injection, 26.0% and 25.8% of NK cells had bound lirilumab in the lirilumab 0.1 and lirilumab 1.0 arms, respectively (Fig. 2C). The frequency of lirilumab-bound Vγ9Vδ2^+^ γδT cells was 12.3% and 8.2% in the lirilumab 0.1 and lirilumab 1.0 arms, respectively. The mean frequency of lirilumab-bound cells was below 6% for CD8^+^ T cells, and below 1% for CD4^+^ T cells in both arms. The frequency of lirilumab-bound immune cells was significantly decreased by C4D1 in the lirilumab 0.1 arm, while maintained in the lirilumab 1.0 arm until the end of study (Fig. 2C).

**Figure 2:**
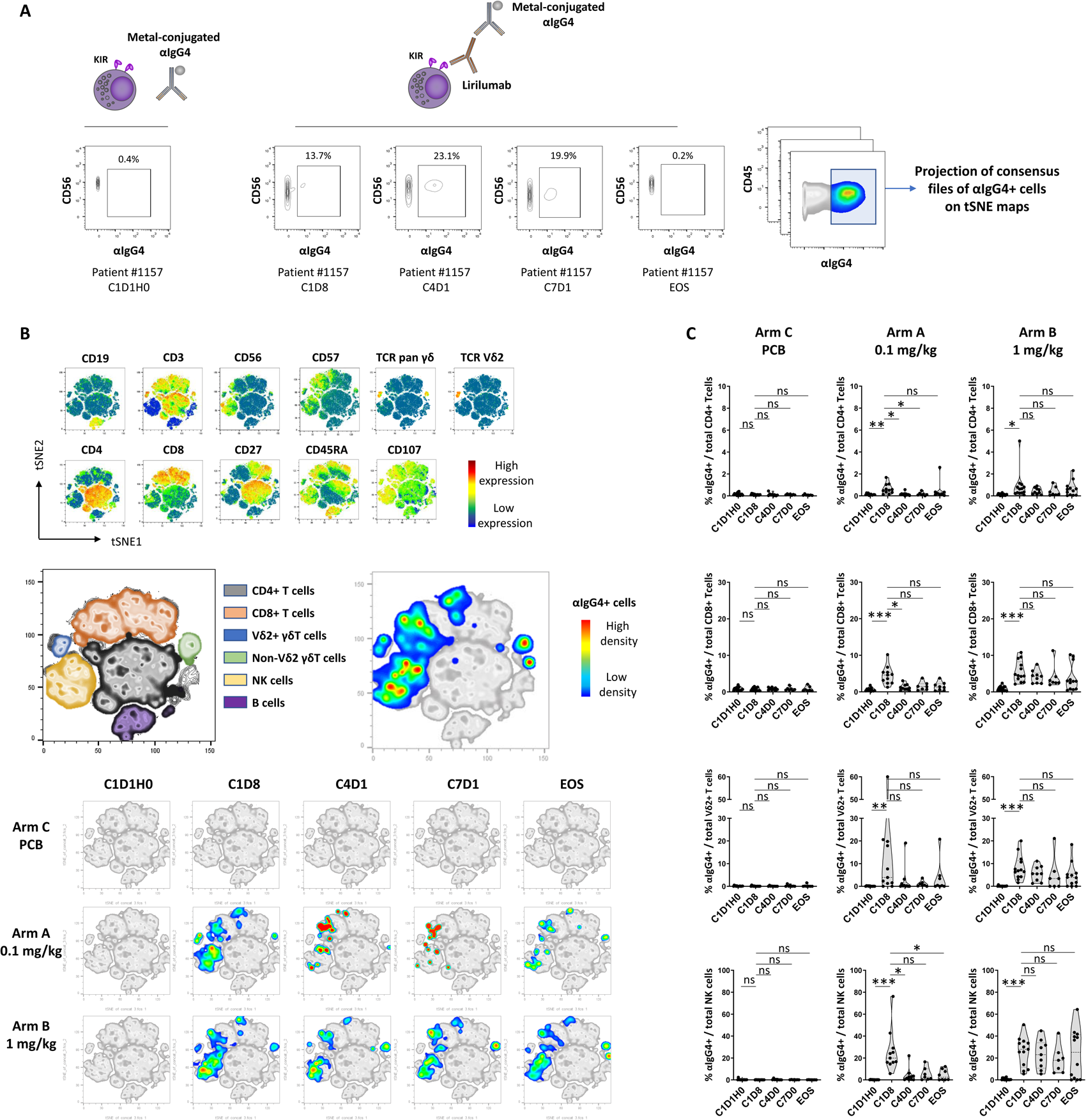
Distribution of lirilumab binding on immune populations. PBMC from patients in arm 0.1 mg/kg (N=12), arm 1.0 mg/kg (N=16) and control arm (N=15) were collected at C1D1H0, C1D8, C4D1, C7D1 and EOS and phenotyped by mass cytometry. A: Lirilumab-bound cells were identified using metal-conjugated an α-IgG4 antibody. B: Consensus files were generated using fixed number of αIgG4+ cells from each .fcs file, and projected on a t-SNE map. C: Frequency of αIgG4+ cells by treatment arm and by time point of blood collection among CD4+ T cells, CD8+ T cells, Vδ2+ γδT cells, and NK cells. Data were analyzed using a Kruskal-Wallis test followed by a Dunn’s post test. \**P* < 0.05; \*\**P* < 0.01; \*\*\**P* < 0.001; ns, nonsignificant. C1D1H0: cycle 1 day 1 hour 0; C1D8: cycle 1 day 8; C4D1: cycle 4 day 1; C7D1: cycle 7 day 1; EOS: end of study.

The impact of lirilumab binding on NK cells was further characterized using dimensionality reduction analysis. Since one of the main hypotheses retained to explain the lack of clinical benefit of anti-KIR therapy in multiple myeloma was the potential interference with NK cell education and development^19,20^, we first focused on NK cell maturation profiles. Lirilumab did not induce any phenotypical shift, even after prolonged administration, except a slight decrease of CD56^bright^ NK cells at end of study (Fig. 3A and Fig. S4A). Besides, we observed a transient decrease of the cluster of KIR^+^ NK cells in the lirilumab 0.1 arm, and a durable decrease of this population in the lirilumab 1.0 arm (Fig. S4B). Analysis of absolute counts of KIR^+^ NK cells in both arms at C1D8 evidenced a weak to drastic decrease in 17 out of 20 patients treated with lirilumab compared with baseline, while this decrease was weak and limited to 3 out of 12 patients in the placebo arm (Fig. 3B), confirming prior observations in multiple myeloma^19^. This decrease of KIR^+^ NK cells was concomitant with a transient decrease at C1D8 of the anti-apoptotic protein Bcl-2 in the lirilumab 1.0 arm, a critical protein NK cell survival.^21^ This decrease affected all the populations of NK cells, independently of lirilumab binding (Fig. 3C), suggesting an indirect mechanism involved in the loss of Bcl-2 consecutive to KIR blockade. Although not significant, a similar trend was also observed in the lirilumab 0.1 arm (Fig. 3C).

**Figure 3:**
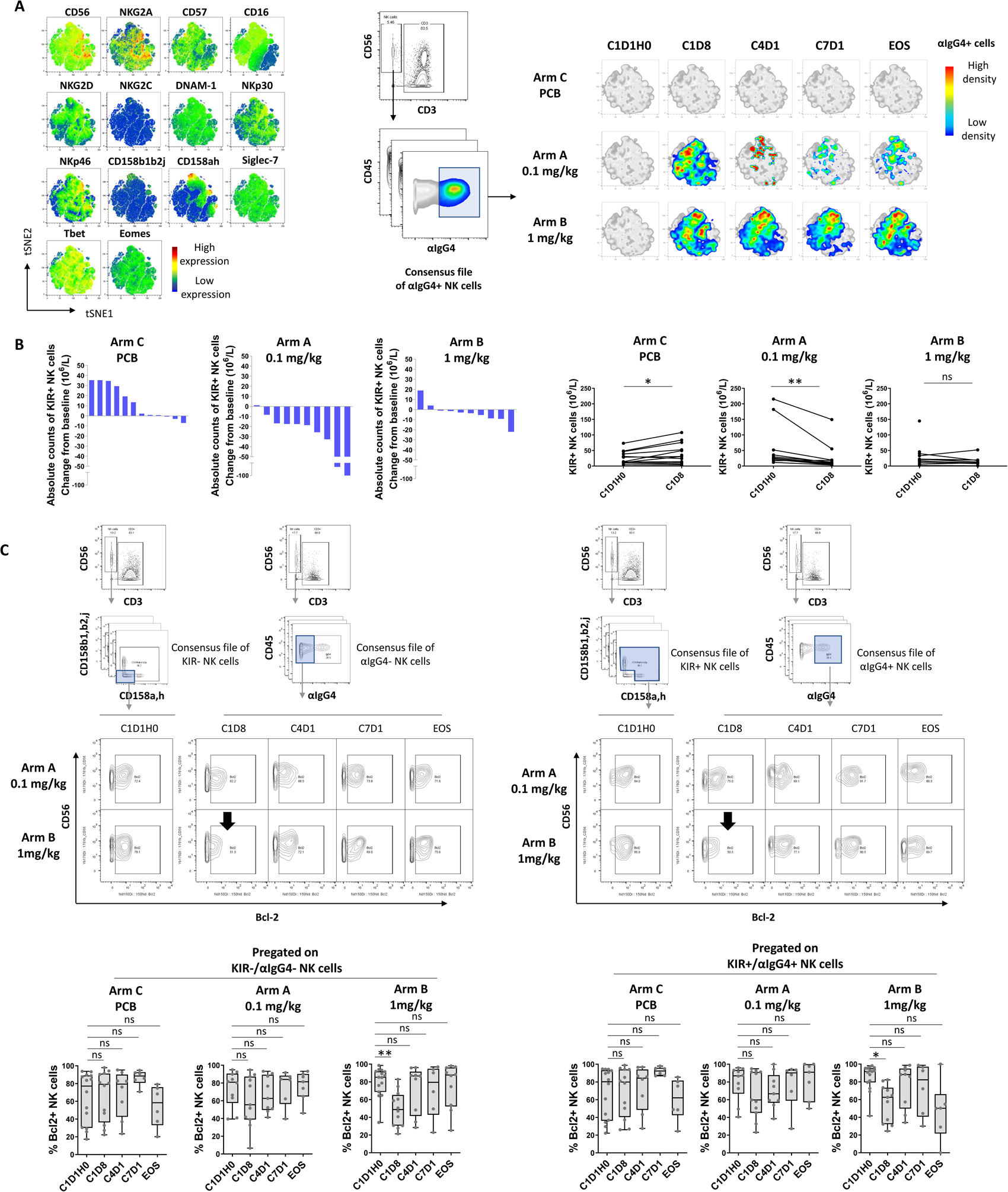
Impact of lirilumab on NK cells. A: Total peripheral NK cells were manually pregated and exported for t-SNE analysis. Consensus files of αIgG4+ NK cells were generated with fixed number of cells for each treatment arm at each time point in order to obtain a representative and balanced view of all patient groups. Consensus files were then projected on t-SNE plots of total NK cells. B: Variations of absolute counts of KIR+ NK cells between baseline and C1D8. C: Bcl-2 expression by treatment arm and by time point of blood collection. C1D1H0 were pre-gated on KIR+ NK cells. C1D8 to EOS were pre-gated on KIR+ cells for placebo arm. C1D8 to EOS were pre-gated on αIgG4+ NK cells for arms A and B. Paired samples were analyzed using a Wilcoxon matched-pairs signed rank test. For multiple comparisons, data were analyzed using a Kruskal-Wallis test followed by a Dunn’s post test, and presented as interquartile ranges, median, and whiskers from minimum to maximum. \**P* < 0.05; \*\**P* < 0.01; ns, nonsignificant. C1D1H0: cycle 1 day 1 hour 0; C1D8: cycle 1 day 8; C4D1: cycle 4 day 1; C7D1: cycle 7 day 1; EOS: end of study; PCB: placebo.

Lirilumab binding on CD8^+^ T cells did not induce any phenotypical shift (Fig. 4A), and did not impact T cell maturation profiles (data not shown). Lirilumab spared naive (CD27^+^ CCR7^+^ CD45RA^+^ CD28^+^), central memory (CD27^+^ CCR7^+^ CD45RA^-^), and effector memory (CD27^-^ CCR7^-^ CD45RA^-^) CD8^+^ T cells, and preferentially bound antigen-experienced CD8^+^ T cells with a terminally differentiated profile (CD27^-^ CCR7^-^ CD45RA^+^ CD28^-^). Most lirilumab-bound CD8^+^ T cells also expressed CD56 and CD16, a phenotypic hallmark of T cells with NK cell-like functions, and enhanced TCR-independent/CD16-dependent degranulation potential.^22,23^ These cells also expressed the long-term immunological memory marker CD57^24^ (Fig. 4A). Upon lirilumab binding, we observed a decreased expression of the activation marker CD69, a key regulator of tumor-specific CD8^+^ T cells differentiation.^25^ This decrease was significant in arm 1.0 at late time points (C7D1), specifically affected lirilumab-bound CD8^+^ T cells, and spared lirilumab-free CD8^+^ T cells (Fig. 4B). We further analyzed the effect of lirilumab binding at early time points on paired samples between C1D8 and C4D1. Lirilumab was found to induce heterogeneous inter-individual responses after binding to CD8^+^ T cells. In 4 out of 8 patients, lirilumab induced up-regulation of the key immune checkpoints CTLA-4, PD-1, and BTLA, which was likely the consequence of a prior activation, as evidenced by higher expression of CD25 (IL-2R), CD28, as well as the inducible co-stimulatory receptors OX40 and 4-1BB (Fig. 4C). In these patients, we also observed an increase of exhausted CD8^+^ T cells characterized by co-expression of PD-1 and TIGIT, two co-inhibitory receptors increased upon T cell activation.^26^

**Figure 4:**
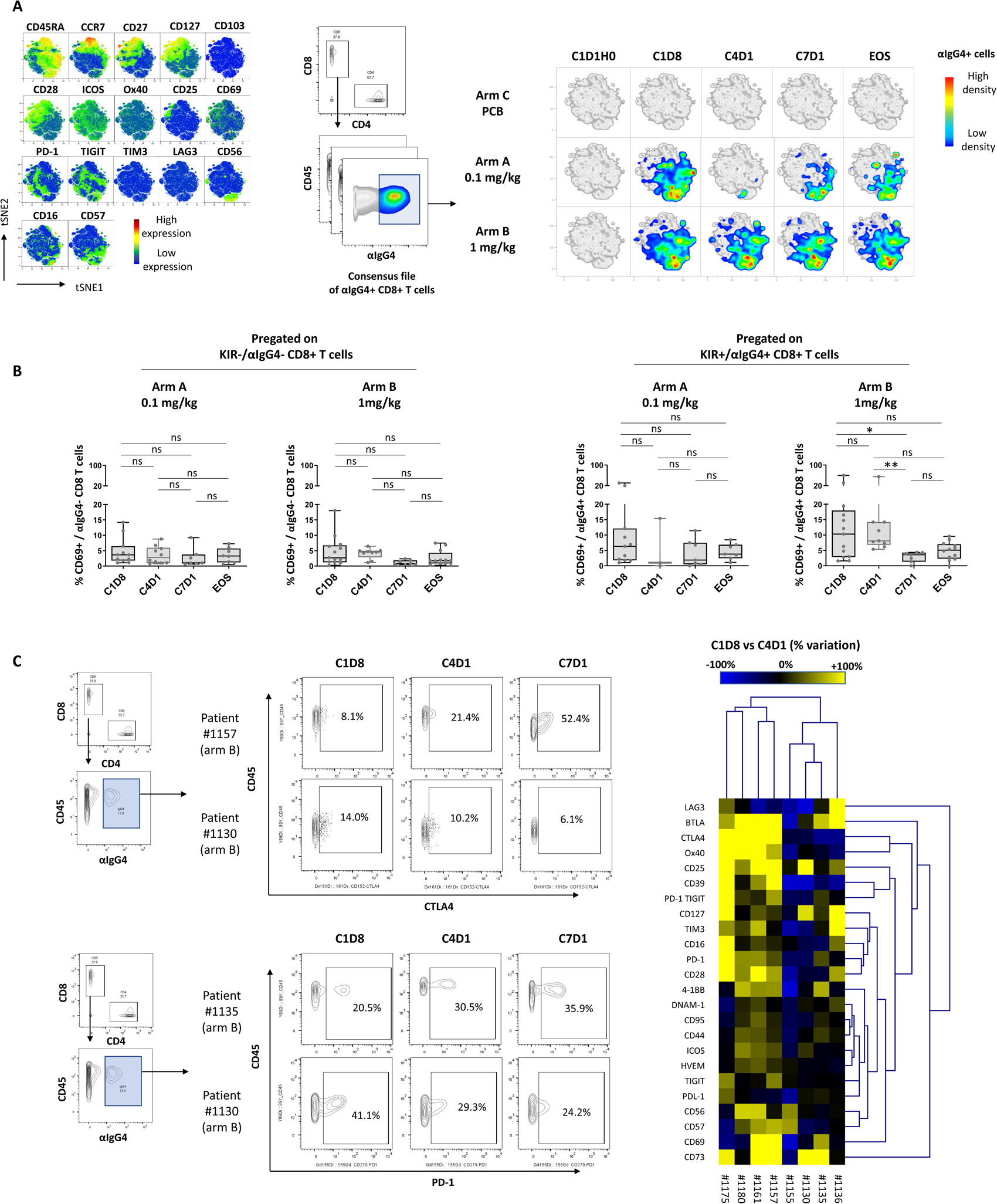
Impact of lirilumab on CD8+ T cells. A: Total peripheral CD8+ T cells were manually pregated and exported for t-SNE analysis. Consensus files of αIgG4+ CD8+ T cells were generated with fixed number cells for each treatment arm at each time point. Consensus files were then projected on t-SNE plots of total CD8+ T cells. B: CD69 expression by treatment arm and by time point of blood collection. C: Variations of T cell triggering receptors, immune checkpoints and activation markers expression between C1D8 and C4D1. Paired samples from patients included in 1.0 arm were analyzed using hierarchical clustering. For multiple comparisons, data were analyzed using a Kruskal-Wallis test followed by a Dunn’s post test, and presented as interquartile ranges, median, and whiskers from minimum to maximum. \**P* < 0.05; \*\**P* < 0.01; ns, nonsignificant. C1D1H0: cycle 1 day 1 hour 0; C1D8: cycle 1 day 8; C4D1: cycle 4 day 1; C7D1: cycle 7 day 1; EOS: end of study.

We next focused on vδ2^+^ γδT cells, a key population of anti-tumor cells with particular relevance in the context of tumors with low mutational burden and high plasticity in MHC-I expression such as AML.^27–30^ Lirilumab mainly bound vδ2^+^ γδT cells with a high cytotoxic potential (CD8^+^ CD56^+^ CD57^+^ CD16^+^) (Fig. 5A). The number of anti-IgG4^+^ γδT cells was insufficient in most patients from arm 0.1 and prevented performing reliable measurements in this group. Upon administration, lirilumab induced a transient decrease of DNAM-1 at C7D1 in arm 1.0, a critical receptor involved in cytotoxicity (Fig. 5B). Besides, we observed a decrease of absolute counts of anti-IgG4^+^ vδ2^+^ γδT cells between C1D8 et C4D1 in arm 1.0 (Fig. 5C). As for NK cells, depletion of lirilumab-bound vδ2^+^ γδT cells was associated with a transient decrease of Bcl-2 at C1D8, affecting vδ2^+^ γδT cells independent of lirilumab binding (Fig. 5B). The effect of lirilumab on Bcl-2 expression was not correlated with treatment schedules, and some patients in arm 0.1 displayed marked decrease of Bcl-2 despite reduced lirilumab dosing. We therefore performed a pooled analysis of both treatment arms according to Bcl-2 expression among patients that relapsed before the end of study. Patients with LFS above the median maintained a high frequency of Bcl-2^+^ vδ2^+^ γδT cells at C4D1, in contrast to patients with LFS below the median (Fig. 5D). Consistently, the frequency of Bcl-2^+^ vδ2^+^ γδT cells was decreased in all patients with LFS<3mo, while the frequency of Bcl-2^+^ vδ2^+^ γδT cells was increased or stable in 8 out of 11 patients with LFS>3mo (Fig. 5E). In line, patients with decreased Bcl-2 expression displayed shorter time to relapse than patients with stable or increased Bcl-2 expression at C1D8 (Fig. 5E). The apparent recovery of Bcl-2 by C4D1 (Fig. 5B) might therefore be artefactual and potentially biased by the fact that most patients with loss of Bcl-2 between C1D0 and C1D8 display LFS<3mo. Taken together, these results highlight a potential link between loss of Bcl-2 in vδ2^+^ γδT cells consecutive to KIR blockade and early relapse in patients treated with lirilumab.

**Figure 5:**
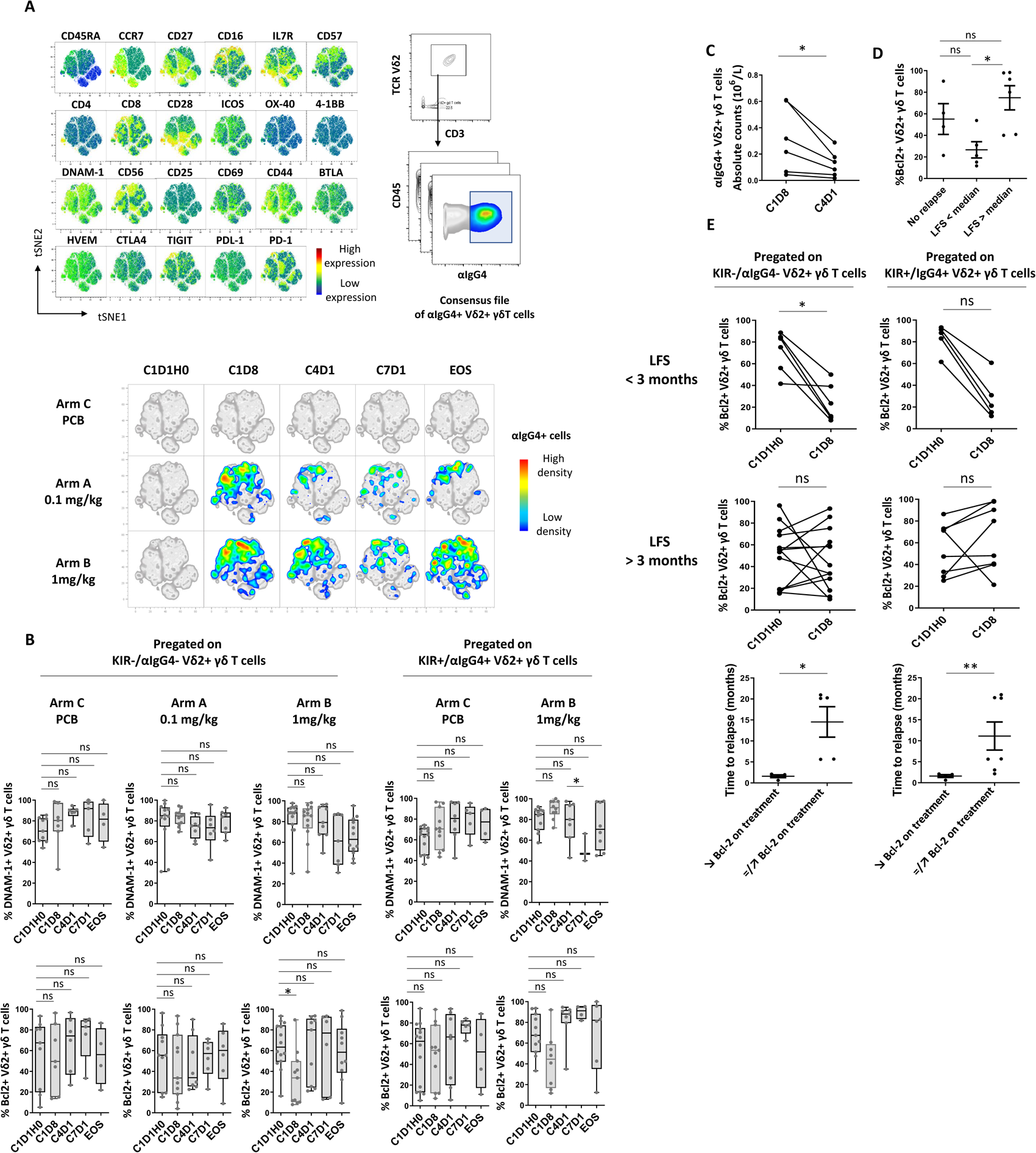
Impact of lirilumab on vδ2+ γδT cells. A: Total peripheral vδ2+ γδT cells were manually pregated and exported for t-SNE analysis. Consensus files of αIgG4+ vδ2+ γδT cells were generated with fixed number cells for each treatment arm at each time point. Consensus files were then projected on t-SNE plots of total vδ2+ γδT cells. B: DNAM-1 and Bcl-2 expression by treatment arm and by time point of blood collection. C1D1H0 were pre-gated on KIR+ NK cells. C1D8 to EOS were pre-gated on KIR+ cells for placebo arm. C1D8 to EOS were pre-gated on αIgG4+ NK cells for arms A and B. C: variations of absolute counts of αIgG4+ vδ2+ γδT cells during the cours of treatment. D: Bcl-2 expression at C1D8 according to LFS. E: Variations of Bcl-2 expression between C1D1H0 and C1D8 according to LFS (upper and middle panels). Time to relapse according to Bcl-2 variations between C1D1H0 and C1D8 (lower panel). For multiple comparisons, data were analyzed using a Kruskal-Wallis test followed by a Dunn’s post test, and presented as interquartile ranges, median, and whiskers from minimum to maximum. Paired samples were analyzed using a Wilcoxon matched-pairs signed rank test. Comparison between two groups were performed using a Mann–Whitney *U* test. \**P* < 0.05; \*\**P* < 0.01; ns, nonsignificant. C1D1H0: cycle 1 day 1 hour 0; C1D8: cycle 1 day 8; C4D1: cycle 4 day 1; C7D1: cycle 7 day 1; EOS: end of study; LFS: leukemia-free survival; PCB: placebo.

## DISCUSSION

The current randomized phase II trial failed to meet its primary objective of demonstrating the efficacy of lirilumab given as maintenance therapy in elderly patients in first CR from AML. The primary endpoint, LFS, did not differ significantly between either of the lirilumab arms and placebo arm, and there were no statistically significant differences in the secondary efficacy endpoints of TTR and OS. However, the lirilumab 0.1mg/kg arm was associated with a non-significant trend toward a higher LFS as compared to the placebo arm (17.6 versus 13.9 months) while an inverse trend was seen in the lirilumab 1mg/kg arm. The latter was stopped early upon a DSMB decision due to the observation of a high rate of early relapse, which translated into a shorter median OS (p=0.058).

We were not able to identify confounding factors which may explain the differences between the 2 lirilumab arms. These include clinical and biological classical AML prognostic factors such as the number of induction courses to achieve a CR, cytogenetics, and ELN genetic risk classification. However, important parameters such as the level of minimal residual disease at inclusion could not be assessed. Doses of 0.1mg/kg of lirilumab were associated with full but transient KIR saturation on NK cells while 1mg/kg dose every 4 weeks led to continuous saturation. However, it must be noted that substantial inter-patient variability was observed and that some patients in the low dose arm actually achieved continuous full KIR saturation.

In order to decipher the mechanism involved in the excess of early relapse observed, we developed mass cytometry panels to track lirilumab-bound cells during the course of treatment, and dissect its impact on immune cell fate. We evidenced a decrease of KIR^+^ NK cell absolute counts following KIR blockade, confirming the results of the phase II trial assessing lirilumab in multiple myeloma.^19^ One hypothesis retained to explain the lack of clinical benefit of anti-KIR therapy in this setting was the potential interference with NK cell maturation homeostasis^20^, the latter being critical in AML as recently evidenced by our group.^5^ We failed to evidence maturation defects following lirilumab administration. Rather, we evidenced a decrease of Bcl-2, a downstream target of IL-15 signaling, involved in mitochondrial apoptosis, autophagy and senescence.^31–33^ The ability of lirilumab to target T cells was notable, confirming prior pre-clinical observations.^18^ Lirilumab preferentially bound antigen-experienced CD8^+^ T cells, leading to a transient decreased activation. The impact of lirilumab on the vδ2^+^ γδT cell compartment was marked by a decreased expression of DNAM-1 and Bcl-2, the latter being associated with a decrease of the absolute counts of KIR^+^ γδT cell, echoing previous work showing that inhibitory KIR (iKIR) expression on human T cells is associated with higher levels of Bcl-2, and that blocking the iKIR-HLA interaction significantly decreased the count of live T cells^34,35^. Similar observations have also been reported in the context of renal cell carcinoma: blocking the interaction of KIR/HLA-Cw4 resulted in the restoration of tumor-induced cell death following activation by interfering with two proximal events of Fas signaling pathway, a sustained c-FLIP-L induction and a decrease in caspase 8 activity.^36^ In line, recent work showed that KIR/HLA interactions extend lifespan of human CD8^+^ T cells.^37^ Although no confirmation of such mechanisms has been reported in γδT cells, we can speculate that similar mechanisms triggering cell death are engaged following KIR blockade on these cells, the latter point requiring dedicated mechanistic studies. Finally, this loss of Bcl-2 was associated with a drastic reduction of time to relapse. Given the strong correlation between Bcl-2 expression by γδT cells, CD8^+^ T cells and NK cells, we cannot exclude a cumulative negative effect of Bcl-2 down-regulation on these three main anti-leukemic populations. However, these results extend the list of arguments in favor of the central role of γδT cells in the control of leukemic blast proliferation.

In conclusion, this study failed to demonstrate that lirilumab can prolong CR duration. In addition, we observed potentially deleterious effects on various immune effectors. These effects - which have not been previously reported - suggest that combined inhibitory effects may have resulted in the impairment of immune surveillance associated with an unexpected high rate of early relapse in the group of patients exposed to prolonged full KIR blockade. The results also confirm the importance of γδT cells in the control of AML cells opening new avenues for future immunotherapeutic strategies.^38,39^

Overall, this study underlies the need to closely monitor the consequences of therapeutic monoclonal antibodies binding on immune populations that appear minor in absolute number, especially when these cells play a key role in tumor clearance. It also warns against the possibility that immune interventions be associated with narrow therapeutic windows.

## Data Availability

All data produced in the present work are contained in the manuscript

## Author contribution statement

N.V. and H.D. designed the research. N.V., H.D., P.A,. A.S.C., A.T., N.D., and D.O. designed the ancillary study. N.V. and A.S.C analyzed the data and drafted the article. N.V., C.R., L.G., B.L., C.E.B, C.P., J.P.L, S.L., E.R., E.R., X.T., Y.H., C.B., B.C., P.R., J.V.M, G.G., J.H.B., M.O.U., M.H., C.P., N.I., and H.D. recruited patients, conducted patient follow-up, and collected clinical data. A.B.A., N.B. and M.S.R. designed the mass cytometry panels and performed the mass cytometry experiments. All authors revised and approved the manuscript.

