## Supplementary material for "Randomized Phase 2 Trial of Lirilumab as maintenance Treatment in Acute Myeloid Leukemia: Results of the EFFIKIR Trial": Sup_material

**Online appendix**

Mass cytometry staining protocol

PBMCs were washed with RPMI 1640 with 10% fetal calf serum (FCS) and incubated in RPMI 1640 with 2% FCS and Pierce Universal Nuclease 25 kU (Thermo Fisher Scientific) diluted at 1:10000 and incubated at 37°C with 5% $CO_2$ for 30 min. Cells were incubated with cisplatin 1 µM to stain dead cells (Standard Biotools). Aspecific epitopes were blocked using 0.5 mg/mL Human Fc Block (BD Biosciences) for 10 minutes at 4°C. Two panels of 35 and 40 antibodies were developed for deep phenotyping of NK cells, $CD4^+$ T cells, $CD8^+$ T cells and $V\delta2^+$ γδ T cells (Table S1). A total of 0.5 to 3 million PBMCs were incubated for 1 hour at 4°C with the extracellular antibodies. Cells were then fixed using 2% PFA. Cells were washed and permeabilized with the Foxp3 Staining Buffer Set (eBioscience) for 30 min at 4°C. Intracellular aspecific epitopes were blocked with 0.5 mg/mL Fc Block for 10 min at 4°C before incubation with the mix of intracellular antibodies for 30 min at 4°C in Foxp3 Staining Buffer. Cells were washed and samples were incubated with intracellular antibodies. Cells were then washed and incubated overnight with 125 nM iridium intercalator (Standard Biotools) in 2% PFA. Finally, cells were diluted in EQ™ Four Element Calibration Beads (Standard Biotools) before acquisition on a Helios instrument (Standard Biotools). Raw .fcs files were manually pretreated using FlowJo v10.8.1. After removal of beads, debris were excluded and cells were gated based on iridium positivity. Dead cells were excluded based on cisplatin positivity.

**Supplementary Table 1:** Mass cytometry panels

Panel 1

| **Antigen** | **Metal** | **Localization** | **Company** |
|---|---|---|---|
| CD45 | 89 Y | Extracellular | Standard Biotools |
| CD3 | 115 In | Extracellular | Immunomonitoring platform |
| CD45RA | 143 Nd | Extracellular | Standard Biotools |
| CD270 (HVEM) | 144 Nd | Extracellular | Standard Biotools |
| CD8α | 146 Nd | Extracellular | Standard Biotools |
| CD278 (ICOS) | 148 Nd | Extracellular | Standard Biotools |
| CD25 (IL-2R) | 149 Sm | Extracellular | Standard Biotools |
| OX40 | 150 Nd | Extracellular | Standard Biotools |
| CD103 | 151 Eu | Extracellular | Standard Biotools |
| CD39 | 152 Sm | Extracellular | Biolegend |
| TIM-3 | 153 Eu | Extracellular | Standard Biotools |
| TIGIT | 154 Sm | Extracellular | Standard Biotools |
| IgG4 | 147 Sm | Extracellular | Biolegend |
| CD279 (PD-1) | 155 Gd | Extracellular | Standard Biotools |
| CD274 (PD-L1) | 156 Gd | Extracellular | Standard Biotools |
| 4-1BB | 158 Gd | Extracellular | Standard Biotools |
| CCR7 | 159 Tb | Extracellular | Standard Biotools |
| CD28 | 160 Gd | Extracellular | Standard Biotools |
| CD272 (BTLA) | 163 Dy | Extracellular | Standard Biotools |
| CD95 | 164 Dy | Extracellular | Standard Biotools |
| CD127 (IL-7R) | 165 Ho | Extracellular | Standard Biotools |
| CD44 | 166 Er | Extracellular | Standard Biotools |
| CD27 | 167 Er | Extracellular | Standard Biotools |
| CD69 | 168 Er | Extracellular | Biolegend |
| TCRVδ2 | 169 Tm | Extracellular | Beckman Coulter |
| CD226 (DNAM-1) | 171 Yb | Extracellular | Standard Biotools |
| CD57 | 172 Yb | Extracellular | Standard Biotools |
| CD73 | 173 Yb | Extracellular | Biolegend |
| CD4 | 174 Yb | Extracellular | Standard Biotools |
| LAG3 | 175 Lu | Extracellular | Standard Biotools |
| CD56 | 176 Yb | Extracellular | Standard Biotools |
| CD16 | 209 Bi | Extracellular | Standard Biotools |
| CD14 | 170 Er | Extracellular | Biolegend |
| CD15 | 170 Er | Extracellular | Biolegend |
| CD13 | 170 Er | Extracellular | Biolegend |
| CD33 | 170 Er | Extracellular | Biolegend |
| CD34 | 170 Er | Extracellular | Biolegend |
| Ki-67 | 141 Pr | Intracellaular | Biolegend |
| Foxp3 | 162 Dy | Intracellaular | Invitrogen |
| CD152 (CTLA-4) | 161 Dy | Intracellaular | Standard Biotools |

Panel 2

| Antigen | Metal | Localization | Company |
|---|---|---|---|
| CD45 | 89 Y | Extracellular | Standard Biotools |
| CD3 | 115 In | Extracellular | Immunomonitoring platform |
| TCRVδ2 | 141 Pr | Extracellular | Beckman coulter |
| CD19 | 142 Nd | Extracellular | Standard Biotools |
| CD45RA | 143 Nd | Extracellular | Standard Biotools |
| CD4 | 145 Nd | Extracellular | Standard Biotools |
| CD8α | 146 Nd | Extracellular | Standard Biotools |
| NKG2C | 147 Sm | Extracellular | Miltenyi |
| SIGLEC 7 | 152 Sm | Extracellular | Biotechne |
| TCRpanγδ | 153 Eu | Extracellular | Beckman coulter |
| CD158b1/b2j | 154 Sm | Extracellular | Beckman coulter |
| CD27 | 155 Gd | Extracellular | Standard Biotools |
| CD96 | 160 Gd | Extracellular | BD |
| NKp46 | 162 Dy | Extracellular | Standard Biotools |
| CD158a/h | 163 Dy | Extracellular | Beckman coulter |
| NKG2A | 165 Ho | Extracellular | Miltenyi |
| NKG2D | 166 Er | Extracellular | Standard Biotools |
| NKp30 | 169 Tm | Extracellular | Beckman coulter |
| DNAM-1 | 171 Yb | Extracellular | Standard Biotools |
| CD57 | 172 Yb | Extracellular | Standard Biotools |
| IgG4 | 174 Yb | Extracellular | Biolegend |
| CD56 | 176 Yb | Extracellular | Standard Biotools |
| CD16 | 209 Bi | Extracellular | Standard Biotools |
| CD14 | 170 Er | Extracellular | Biolegend |
| CD15 | 170 Er | Extracellular | Biolegend |
| CD13 | 170 Er | Extracellular | Biolegend |
| CD34 | 170 Er | Extracellular | Biolegend |
| CD33 | 170 Er | Extracellular | Biolegend |
| Bcl-2 | 150 Nd | Intracellular | Biolegend |
| Bcl-XL | 158 Gd | Intracellular | Biolegend |
| KI 67 | 159 Tb | Intracellular | Biolegend |
| CD107α | 151 Eu | Intracellular | Standard Biotools |
| EOMES | 149 Sm | Intracellular | BD Pharmigen |
| Tbet | 161 Dy | Intracellular | Standard Biotools |
| Granzyme B | 173 Yb | Intracellular | Standard Biotools |

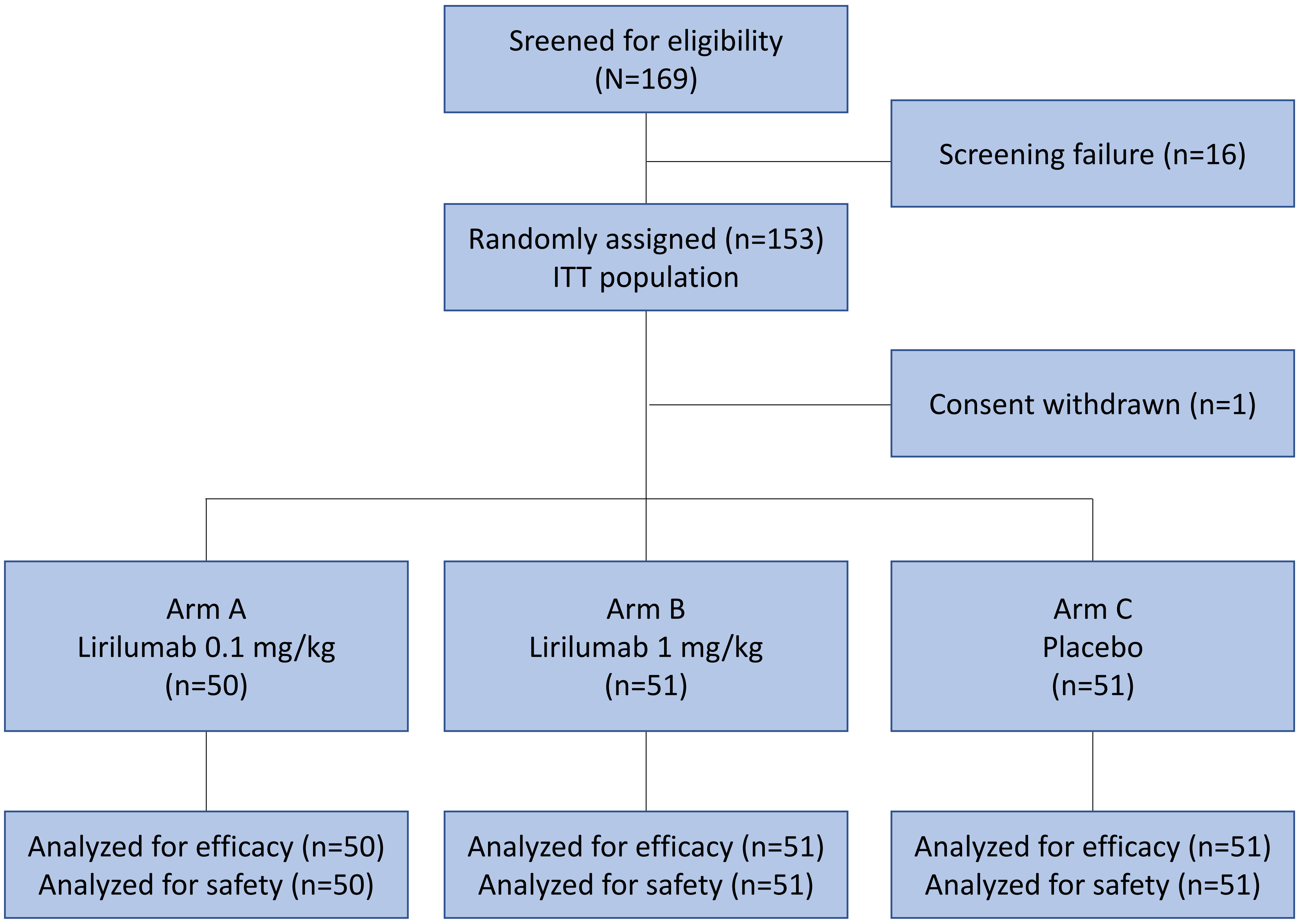


**Supplementary Figure 1**: Consort diagram.

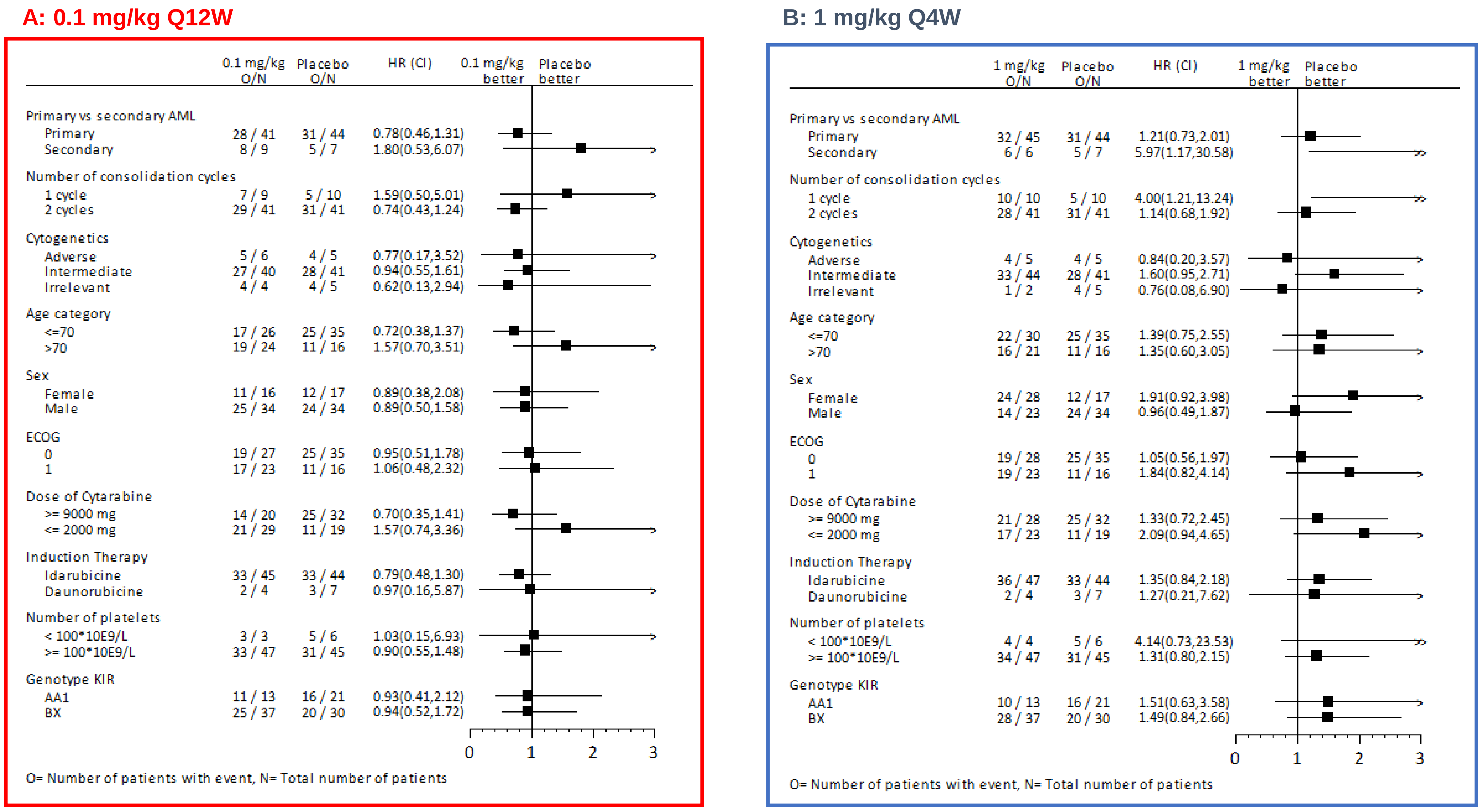


**Supplementary Figure 2**: Factors affecting LFS (Forest Plot analysis). A: lirilumab 0.1 mg/kg arm versus placebo; B: lirilumab 1.0 mg/kg arm versus placebo.

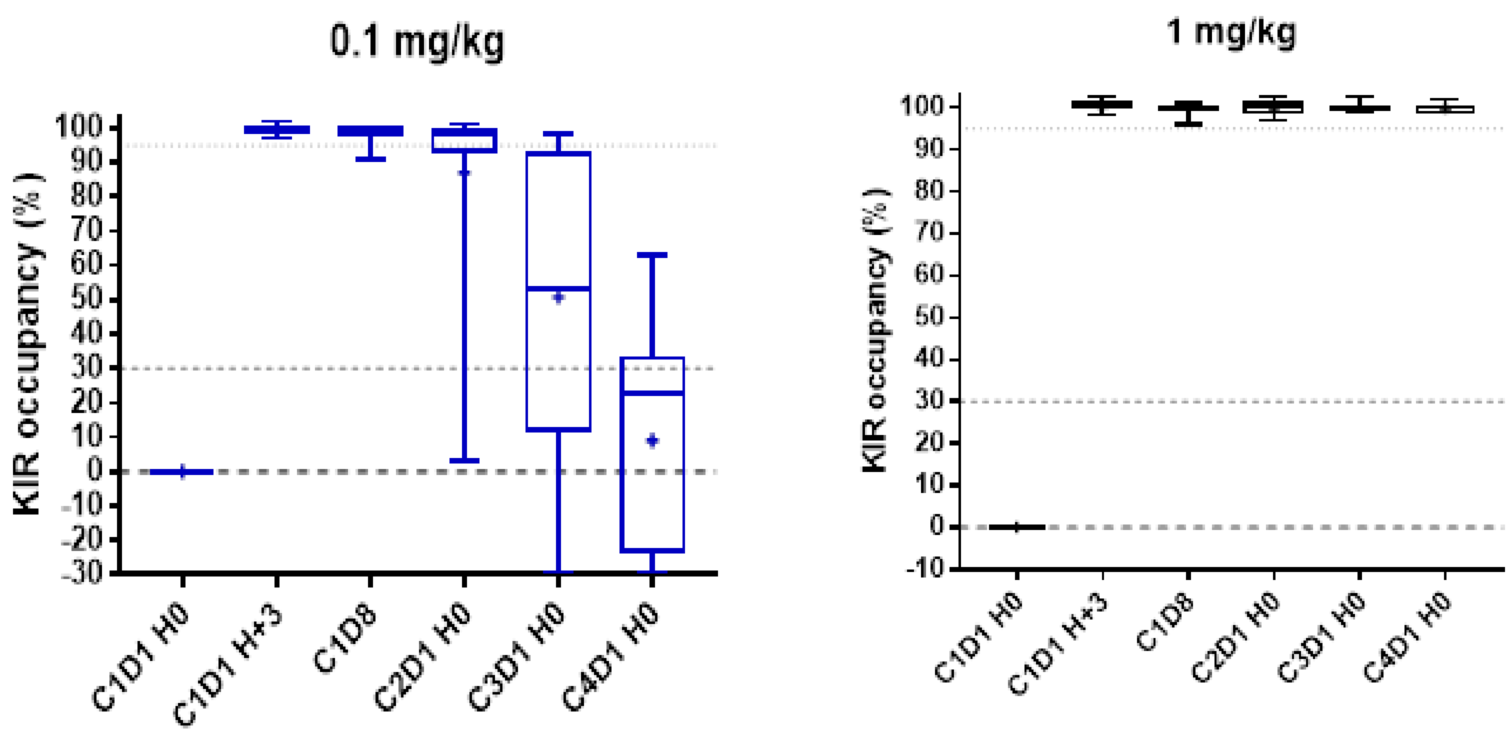


**Supplementary Figure 3.** KIR Occupancy According to Lirilumab Dose. KIR occupancy was assessed for the first 60 patients randomized at screening, C1 at H0 (pre-dose) and H+3; C1 Day 8; C2, C3 and C4 at H0. KIR saturation on peripheral-blood NK cells was assessed on whole blood by flow cytometry. KIR occupancy evaluation was based on the detection of free KIR on NK cells at each timepoint in comparison with the amount of KIR expressed on NK cells before dosing. LLOQ: lower limit of quantification.

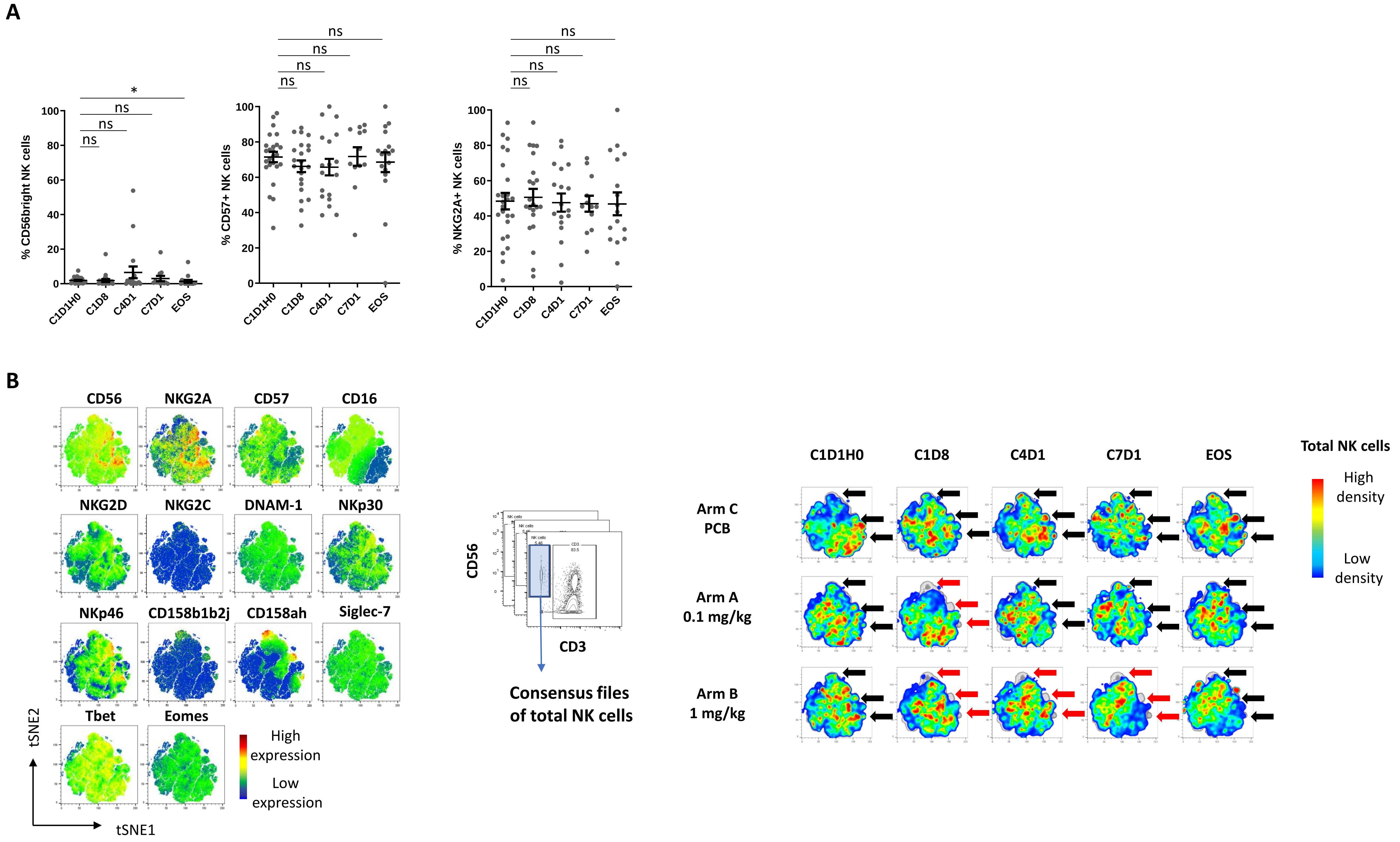


**Supplementary figure 4: Impact of lirilumab on NK cell maturation.** A: NK cell expression CD56, CD57 and NKG2A by time point of blood collection (pooled analysis of arms 1.0 and 0.1). B: Total peripheral NK cells were manually pregated and exported for t-SNE analysis. Consensus files of NK cells were generated with fixed number cells for each treatment arm at each time point. Arrows indicate KIR+ clusters; red arrows indicate decreased density of KIR+ clusters.
